# Anemia in Pregnant Women as a Cultural Phenomenon: A Literature Review

**DOI:** 10.1101/2023.12.22.23300423

**Authors:** Christine Aden, Moses Glorino Rumambo Pandin, Nursalam Nursalam

**Affiliations:** Doctoral Program Student from The Faculty of Nursing, Universitas Airlangga, Jalan Dr. Ir. H. Soekarno, Mulyorejo, Kec. Mulyorejo, Surabaya, East Java 60115 2; Faculty of Humanities Universitas Airlangga, Jalan Dr. Ir. H. Soekarno, Mulyorejo, Kec. Mulyorejo, Surabaya, East Java 60115

**Keywords:** Anemia, Pregnant women, Culture

## Abstract

**Background:** Explores how restrictions on food intake during pregnancy, due to cultural taboos, can lead to anemia. The study aims to provide an overview of prohibited food types, the supportive environment, the impact on pregnancy, and interventions that can be implemented to overcome anemia.

**Method:** The authors conducted a literature review by searching for articles on five databases, namely Science Direct, Springer Link, Pubmed, Sage, and Scopus. They also searched for suitable articles from other references, mainly journal articles published in the last five years (2019-2023).

**Results:** In total, there were 20 reviewed explaining the types of food prohibited during pregnancy and the reasons for this as well as support from family, religious figures, mothers, and in-laws. Overcoming anemia in pregnant women with a cultural approach resulted in meaningful results, with the intervention carried out for around three months. The use of picture books that are appropriate to the cultural context and local wisdom increases information about the importance of iron for pregnant women, increases the frequency of eating, the number of iron tablet intakes, the amount of food intake containing iron each day, and increases the baby’s birth weight.

**Conclusion:** Anemia caused by cultural taboos on certain foods during pregnancy can be prevented and treated with local wisdom intervention approaches.

## BACKGROUND

Anemia in pregnancy is a major nutritional problem in the world. Anemia is experienced by 37% (37 million) pregnant women (WHO, 2023) and around 20% of maternal deaths in the world are caused by anemia. Teshome et al., 2020 state that anemia is a determinant of low body weight, premature birth, and infant death. As per the 2020 global nutrition report, 194 countries have agreed to reduce cases of anemia by 50% among women of childbearing age by 2025 (Muthuraj et al., 2023).

Every pregnant woman needs good food intake for her health and that of the fetus during pregnancy. (Asim et al., 2021). Socio-cultural barriers often cause inadequate nutritional intake in pregnant women (Mohammed et al., 2019). Cultural beliefs, such as food taboos, refer to foods that pregnant women or communities avoid for cultural reasons (Anggraini et al., 2023). Restricting pregnant women from consuming food can cause anemia (Laisser et al., 2022). Inadequate nutritional intake during pregnancy due to taboo patterns can cause pregnant women to experience malnutrition (Anggraini et al., 2023).

Workneh et al (2022) stated that experts previously explained that family members and pregnant women already know the good varied diet and different food types to improve the health of the mother and fetus. However, access to nutritious food and cultural and religious perspectives regarding food intake can be obstacles (Dalaba et al., 2021). Restricted access to healthy eating patterns due to culture, religion, and hygiene can cause pregnant women to abstain from nutritious food (Romulandi et al., 2023). Additionally, providing unscientific and dangerous nutritional information to pregnant women can result in the dismissal of counseling (Alehegn et al., 2021).

Families have inherited the experience of caring for pregnant women to care for pregnant women in the family (Darmawati et al., 2022). Fathers, mothers, in-laws, and traditional leaders teach pregnant women to obey existing cultural norms to protect them from various obstacles and dangers during pregnancy and childbirth (Maycondo et al., 2022). Half of pregnant women in India live with their mothers-in-law, and they make important decisions regarding the condition of their pregnancy under the guidance of their mother-in-law (Varghese et al., 2019).

The following literature study will review the cultural values that pregnant women adopt to abstain from certain foods, the impact of anemia that accompanies it, and the programs that can help overcome it. Previous literature studies did not explain the supportive environment that pregnant women need to follow food habits during their pregnancy. This research aims to explain the types of taboo foods that should be avoided and interventions that can be implemented to overcome anemia by adapting to culture.

## Method

The guidelines for carrying out a literature review follow the Preferred Reporting Items for Systematic Review and Meta-Analysis (PRISMA). The article search strategy included using appropriate keywords such as “culture”, “support”, “anemia”, and “pregnancy”, as well as searching articles through databases such as Science Direct, Springer Link, Pubmed, Sage, and Scopus. The journal articles selected for this study are from the last five years (2019- 2023). The researchers also used the PRISMA flow diagram to obtain research articles that meet the requirements. The PRISMA flow diagram is shown below:

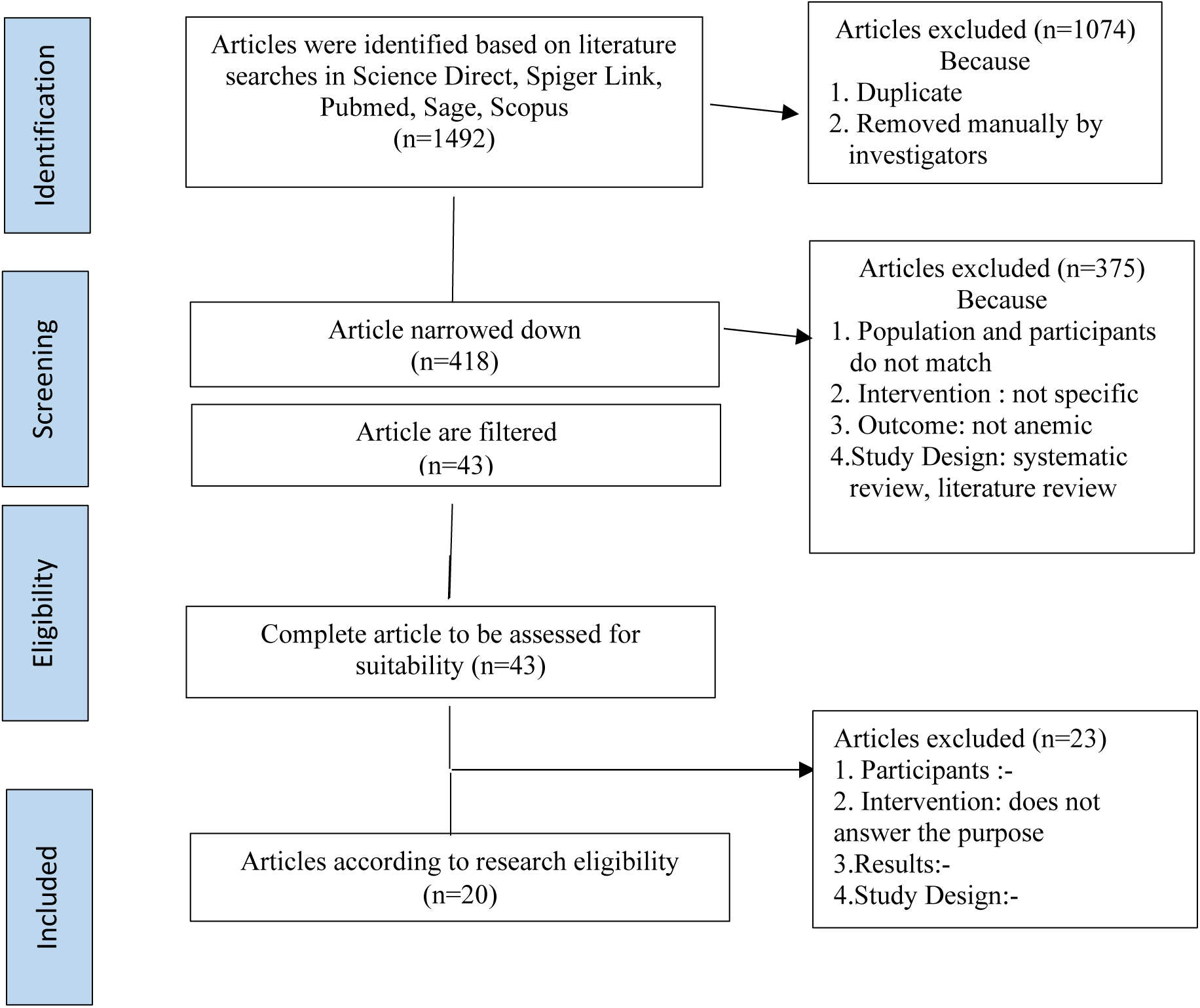

## Results

The author found 1492 articles from 5 databases Science Direct, Springer Link, Pubmed, Sage, and Scopus. After selecting for duplication via the Rayyan application and manually and based on PICO, namely population, intervention, comparison, and results, 20 articles were obtained that followed the research objectives. The results of articles showed that 6 articles explained the types of foods that are prohibited during pregnancy and the reasons. as well as the role of family, religious figures, mothers, and in-laws. The article was explained by Workneh et al (2023), Asim et al (2022); Amare et al (2022); Maykondo et al (2022); Dalaba et al (2021), Ramulondi et al (2021), and Alehegn et al (2019). Some articles examine the opinions of traditional leaders, mothers’ perspectives, and husbands’ perceptions of anemia which are explained by 2 articles by Darmawati et al (2020) and 1 article by Darmawati et al (2022). The relationship between anemia and various causal factors is reviewed in 5 articles by Rekha et al (2019); Mohammed et al (2019); Teshome et al (2020); Agbozo et al (2020); Muthuraj et al (2023). Detection of anemia and malnutrition and interventions to treat anemia are described in 4 articles by Angraini et al (20230, Citrakesumasari et al (2020), Nahrisah et al (2020), Papandreou et al (2023), and 1 article on the impact of anemia in pregnant women on babies born is explained by Sah et al (2022). The results of the article review can be seen in 1 below:

**Table 1.**
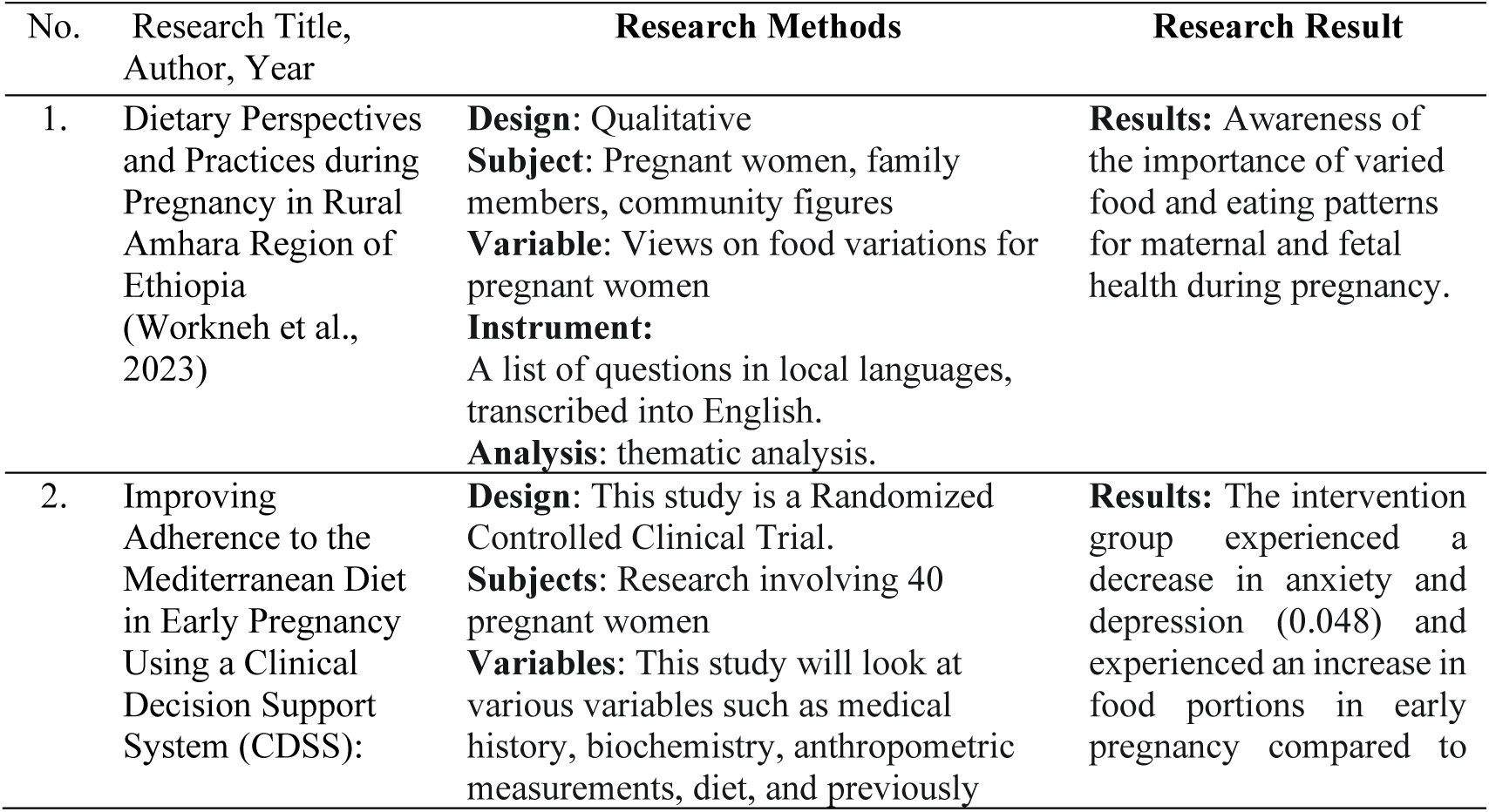
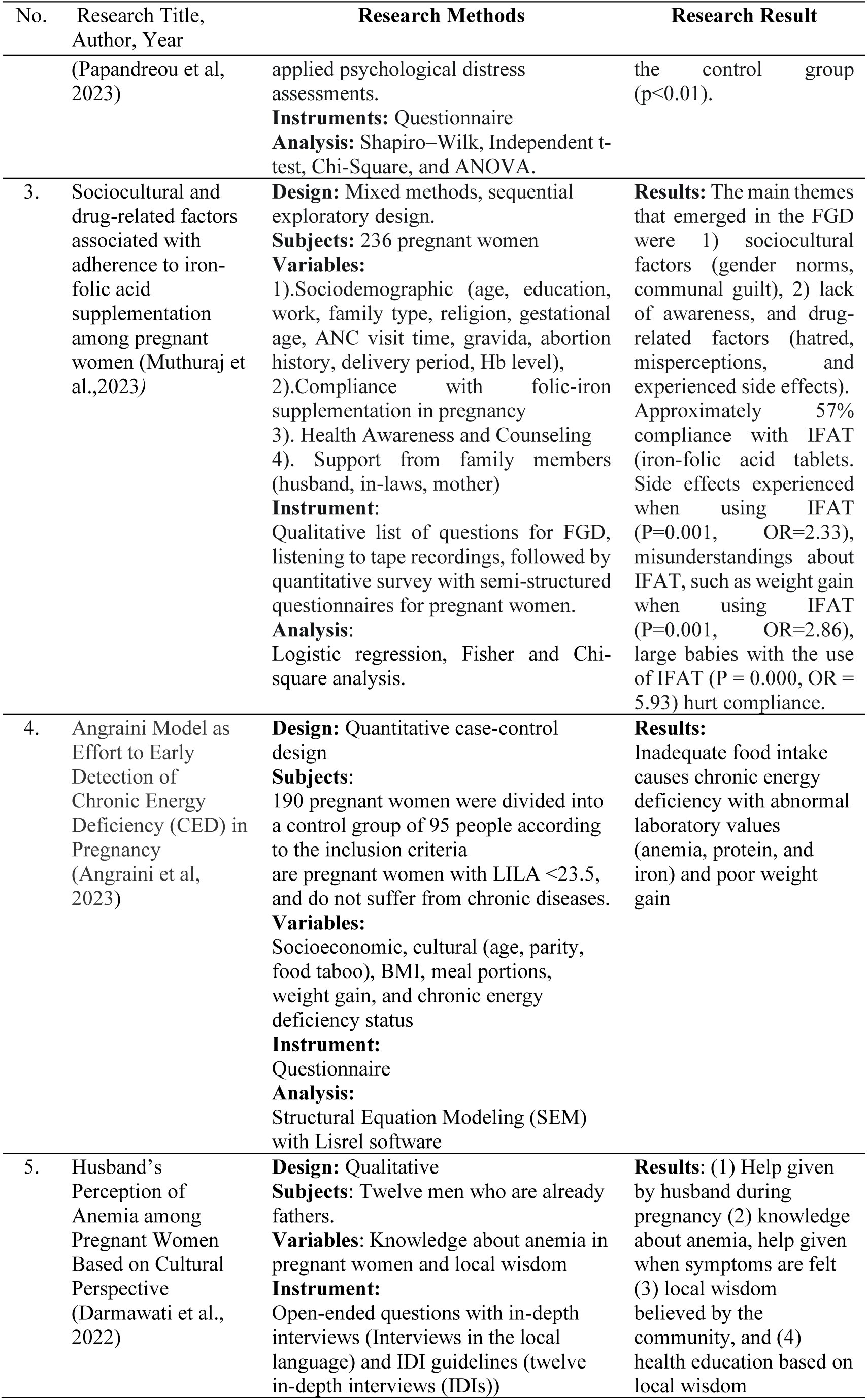
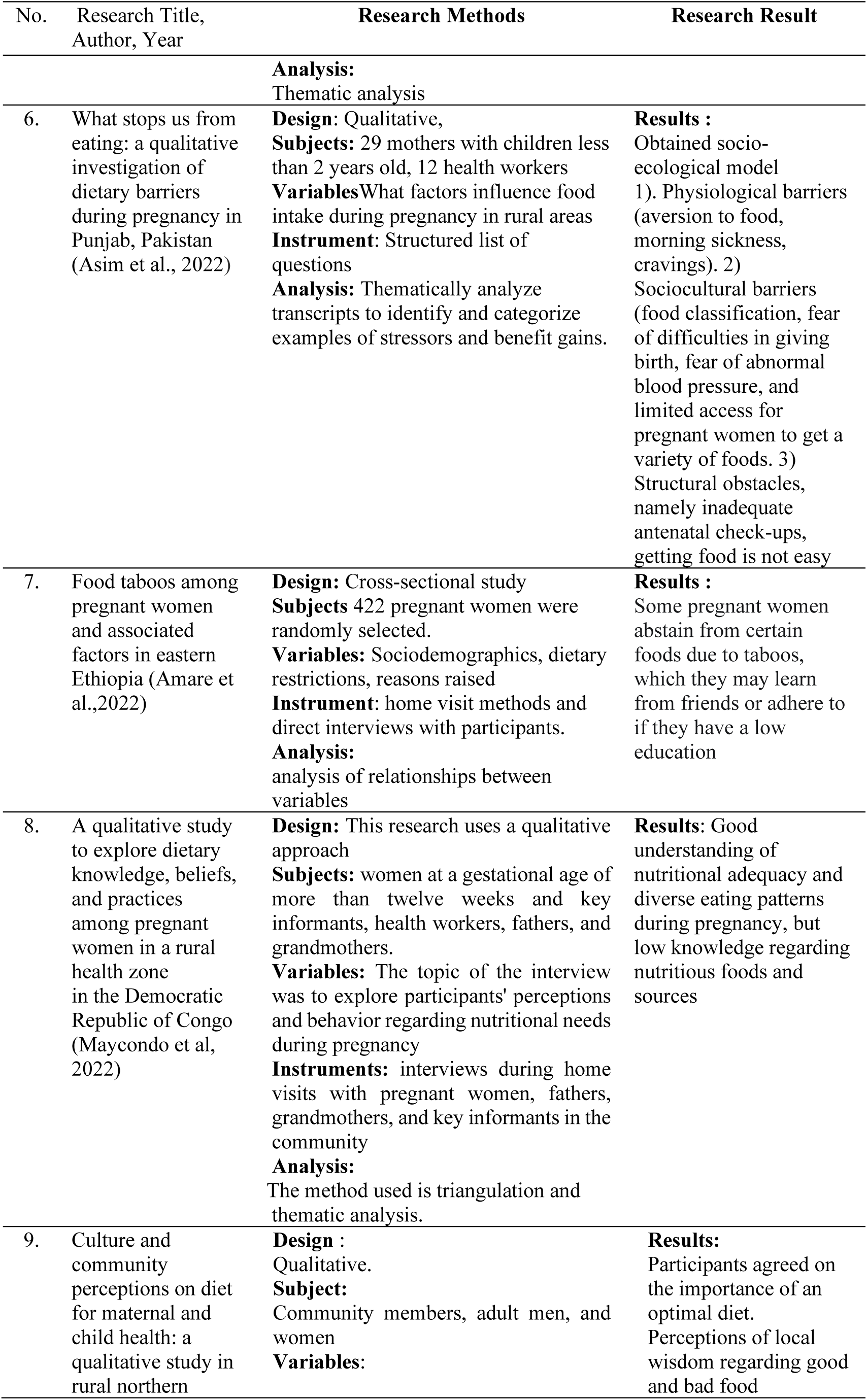
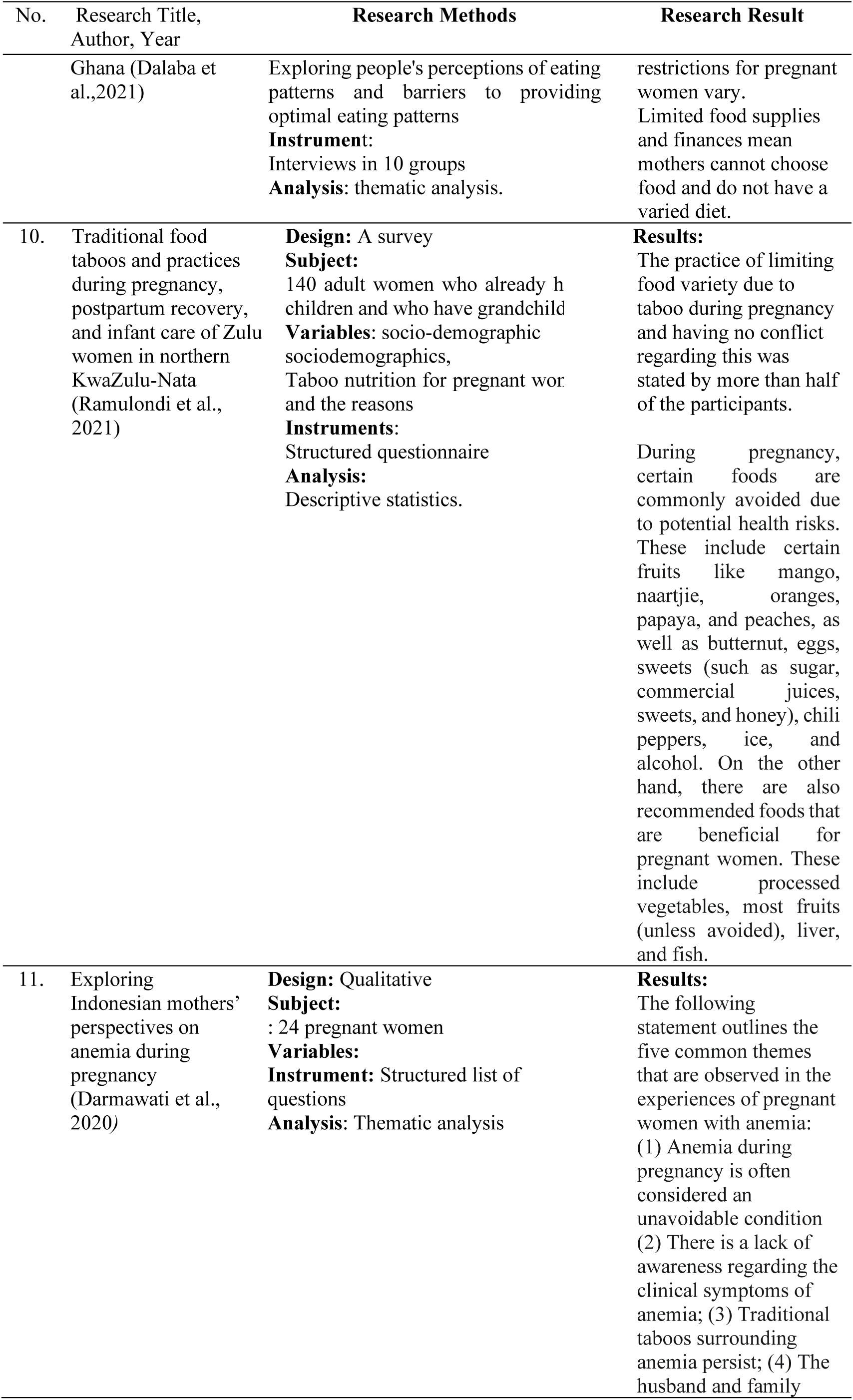
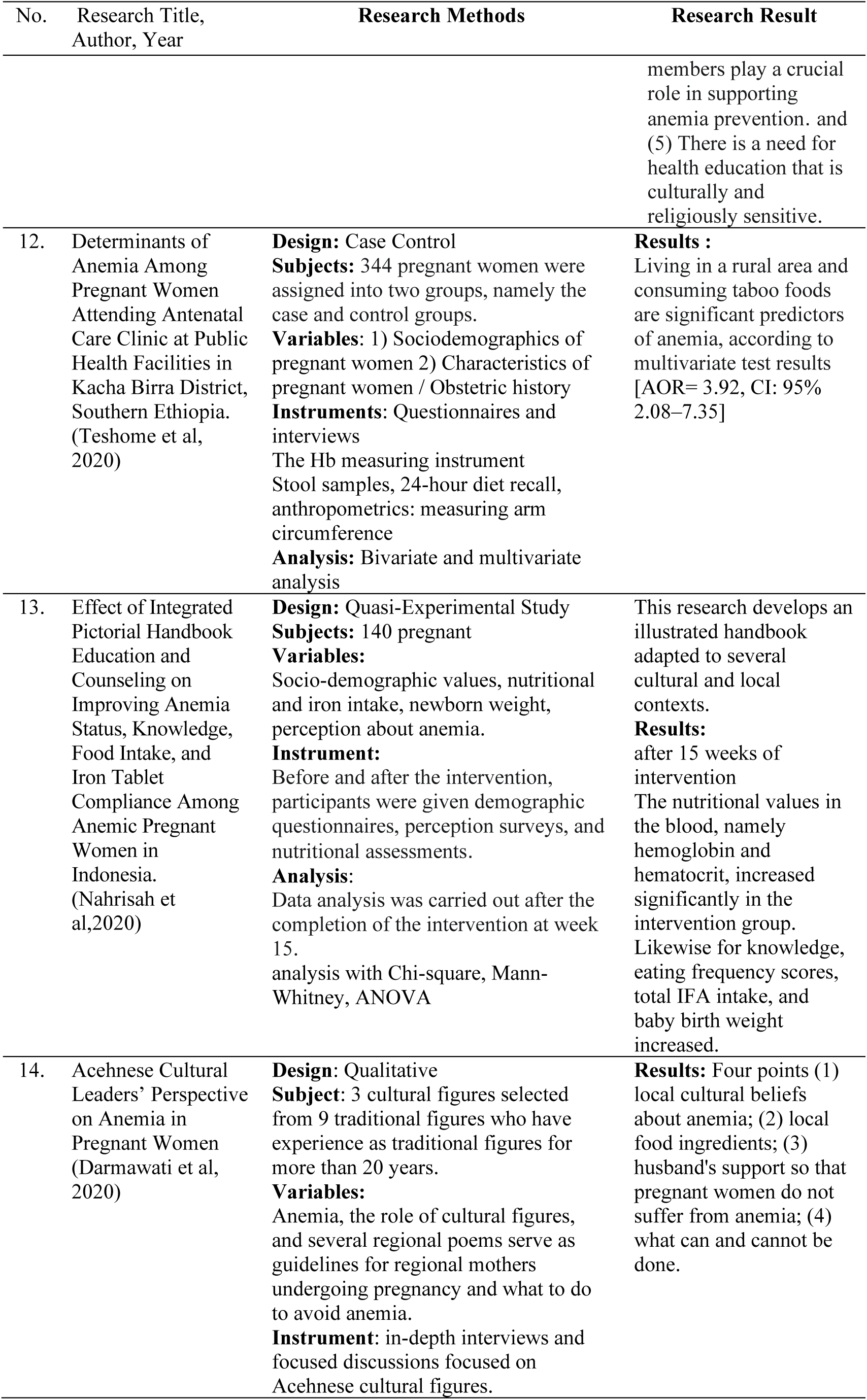
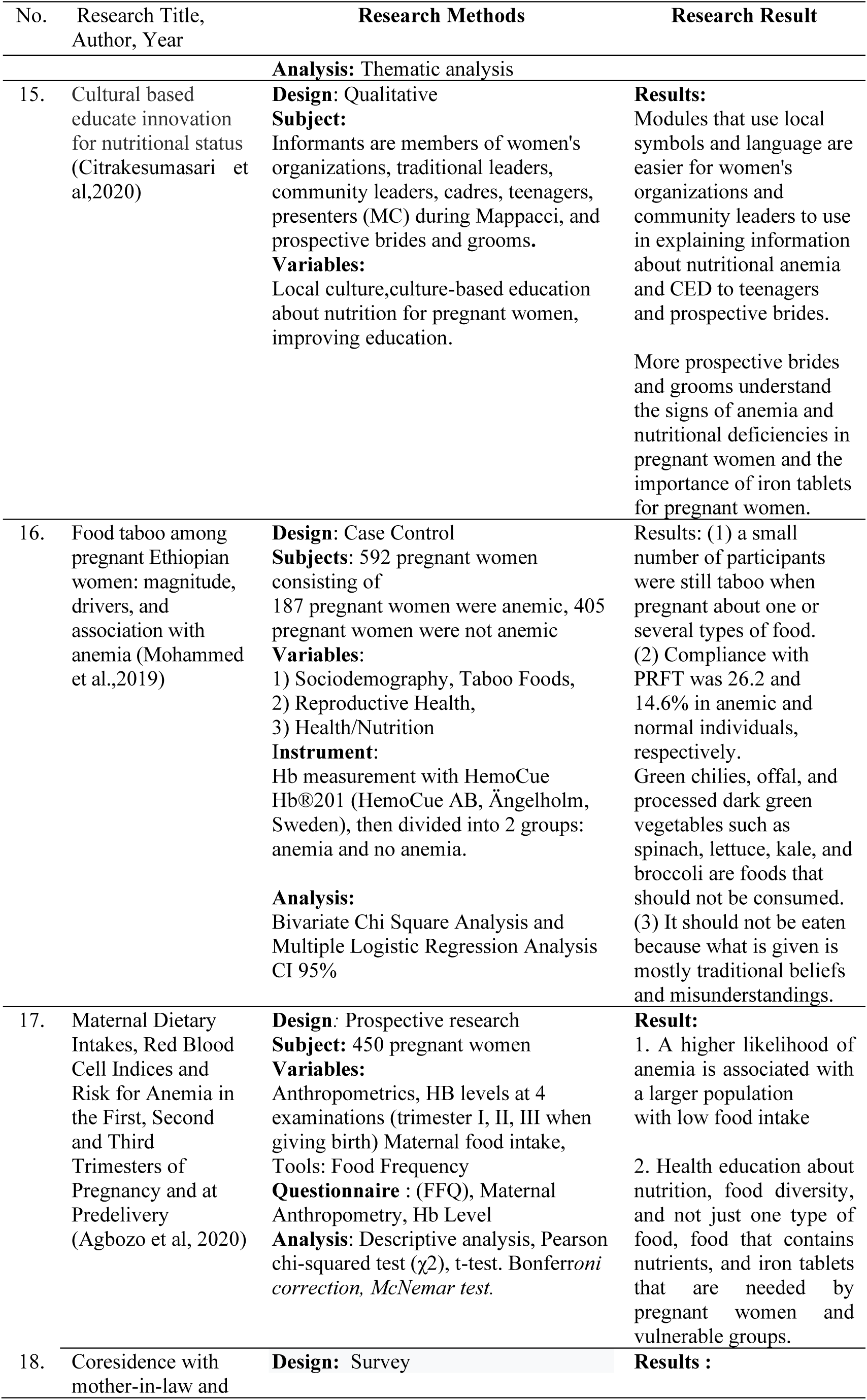
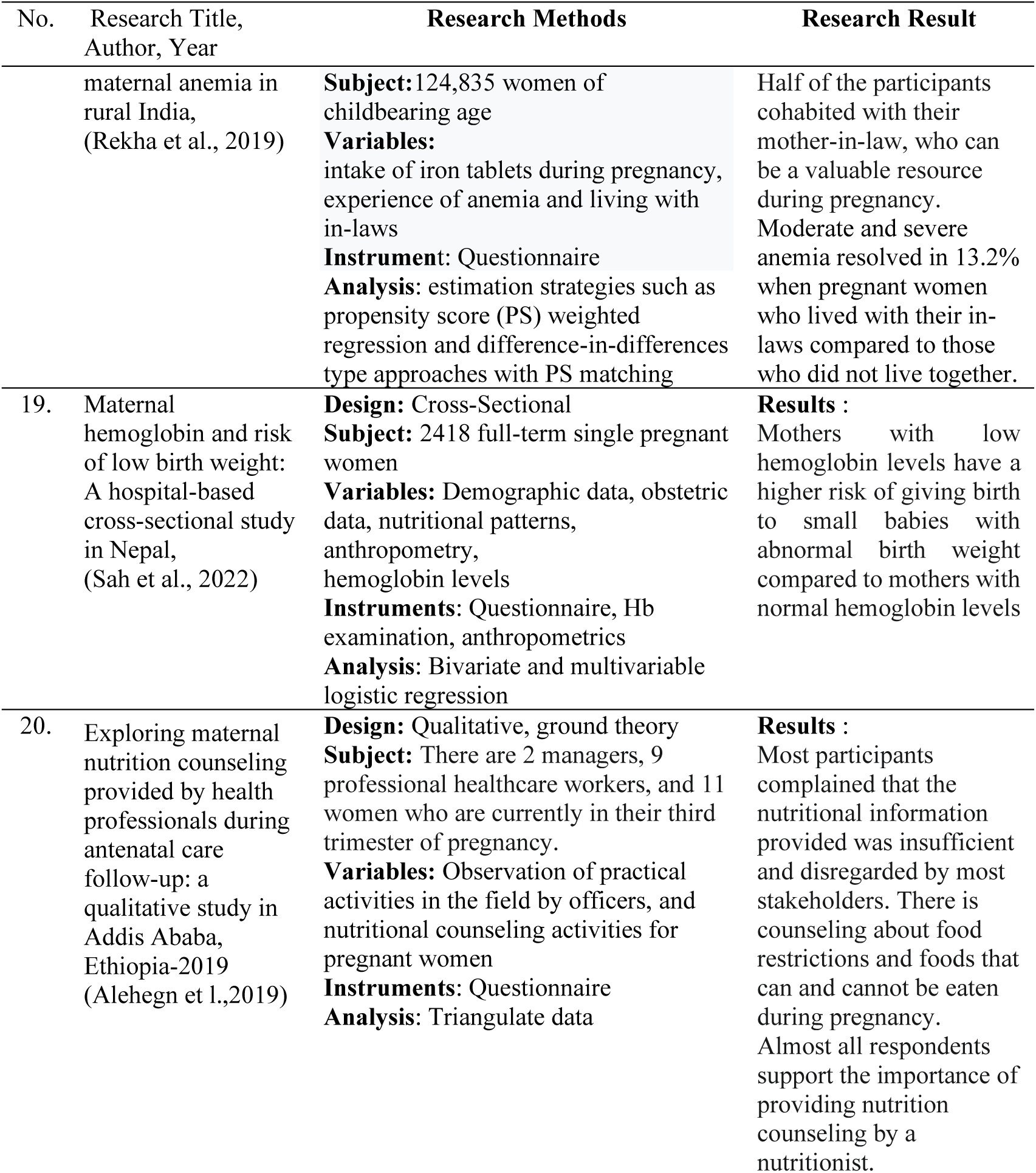
Characteristics of The Studies Reviewed.

## Discussion

Food restrictions in Pakistan are grouped into hot foods, cold foods, and hard foods. The closest family, especially the shaman and in-laws, always reminds me. Pregnant women after 12 weeks of pregnancy or after the symptoms of nausea and vomiting have passed are recommended to eat foods in the cold group because, in the early period of pregnancy, the body is in a hot state. On the other hand, when entering the third trimester at the end of pregnancy, pregnant women are advised to eat foods in the hot group because it accelerates delivery.

During pregnancy, mothers are not advised to consume hard foods, because it can cause the mother’s stomach to bloat and disrupt the condition of the pregnancy (Asim et al., 2022).

Dietary restrictions on animal protein include meat, milk, eggs, and certain fish in Ethiopia and their effects on the mother and future baby (Amare et al, 2022; Workneh et al., 2023). Dark green vegetables such as kale, spinach, broccoli, and lettuce are the foods that are most avoided, as are green chilies, and offal (Mohammed et al., 2019). Pregnant women stated that food taboos in the Republic of Congo were specifically reminded by parents, especially fathers, mothers, and traditional elders and grandmothers who played an important role in regulating the food that pregnant women could consume (Maycondo et al, 2022).

Specifically in Ghana, apart from animal products that are taboo, mushrooms, vegetables, and red fruit are taboo for pregnant women (Dalaba et al, 2021). In contrast to Pakistan, Ethiopia, and Ghana, in Zulu, the detailed food assortment of animal products and vegetable products also includes yellow vegetables and fruit. The impact that occurs on the mother, such as problems during pregnancy and childbirth and on the baby after birth, will experience pain and defects at birth.

The family helps pregnant women learn how to take care of themselves during pregnancy (Darmawati et al., 2022). Fathers, mothers, in-laws, and traditional leaders teach pregnant women to comply with existing cultural norms so that they are protected from various obstacles and dangers during pregnancy and childbirth (Maycondo et al., 2022). Amare et al, 2022 stated that in eastern Ethiopia half (48%) of pregnant women reported having dietary restrictions during pregnancy and having dietary restrictions during pregnancy. Sources of information were family (in-laws), neighbors, community, health workers, and television/radio (OR 3.58; 95%, CI 1.89, 6.83), more (39.2%) friends of pregnant women avoided food during pregnancy (OR 1.91;95%, CI 1.22, 2.99)

The possibility of anemia occurring is 4.21 times greater in pregnant women with a low variety of food types compared to pregnant women with a high variety of food types (Teshome et al., 2020). Agbozo et al (2020) confirmed that the risk of anemia increases 3-4 times during pregnancy. When surveyed, one-fifth of pregnant women reported avoiding certain foods due to cultural taboos (Muhammad et al, 2019). Food restrictions due to taboos are a predictor of anemia. Anemia is 3.9 times more common in pregnant women who observe restrictions due to cultural practices compared to pregnant women who consume various types of food without restrictions (Teshome et al, 2020).

Abstinence from food for a long time can cause Chronic Energy Deficiency (CED) due to a low intake of food containing protein, carbohydrates, fat, and iron (Anggraini et al, 2023). Pregnant women with low iron levels or hemoglobin have a higher risk of abnormal birth weight for their babies compared to women with normal hemoglobin levels (Sah et al, 2022).

All the research results above are about field facts about pregnant women’s compliance with the culture of abstinence from food and are strengthened by information from traditional leaders and supervision from fathers, mothers, mothers-in-law, and husbands as well as fears of the dangers that will be experienced by mothers and babies who will be born. Belief in culture cannot be easily eliminated, so interventions related to food taboos can be carried out with religious-based health counseling (Darmawati et al, 2020) because religious beliefs that fasting for pregnant women is a spiritual practice so as not to harm pregnancy (Workneh et al, 2023). Providing information about local food that contains iron (Darmawati et al., 2020) as well as health education based on local wisdom (Darmawati et al., 2022). Papandreou et al., 2023 stated that the group of pregnant women were more compliant (p<0.01) when given a local food diet in early pregnancy and experienced an increase in nutritional status. Anxiety and depression decreased among pregnant women who received local traditional food menus (p=0.048), compared to the control group.

Overcoming anemia in pregnant women with a cultural approach turns out to get meaningful results, Darmawati et al, 2020, 2022; Papandreau et al., 2023; Nahrisah et al., 2020; Citrakesumasari et al, 2020 with intervention carried out for around 15 weeks. The use of picture books that were appropriate to the cultural context and local wisdom in the control group increased hemoglobin and hematocrit levels, increased knowledge, eating frequency scores, number of iron tablets intake, and total intake of other substances daily iron from food and the baby’s birth weight (Nahrisah et al., 2020). Likewise, the use of modules with symbols and local language that are per local wisdom can increase the bride and groom’s knowledge about the signs and risks of anemia, nutrition during pregnancy, and CED (Citrakesumasari et al., 2020).

## Conclusion

Food restrictions during pregnancy come from animal products and plant products. The culture of abstinence is supported by traditional elders and family, namely the father, mother, in- laws, and husband. Anemia and prolonged food intake can cause Chronic Energy Deficiency (CED) in mothers and babies with low birth weight. Interventions to prevent anemia can be carried out by providing health education using local cultural and religious approaches and providing food from local food sources.

## Data Availability

All data produced in the present work are contained in the manuscript

## Acknowledgements

We thank all the lecturers at Airlangga University and friends from Class Eight who have supported the writing of this article.

## References

Teshome, MS., Meskel, DH., Wondafrash, B. (2020). Determinants of Anemia Among Pregnant Women Attending Antenatal Care Clinic at Public Health Facilities in Kacha Birra District, Southern Ethiopia, Journal of Multidisciplinary Healthcare 2020:13 1007–1015, PMID: 33061406 PMCID: PMC7522419 doi: 10.2147/JMDH.S259882

Muthuraj, LP., Kandasamy, S., Subbiah, P., Sibqathulla, MJ., Velappan, LK., Gopal, M., Ramya, JE., Jayaraman, Y., and Kalyanaraman, S. (2023). Sociocultural and drug-related factors associated with adherence to iron– folic acid supplementation among pregnant women. J Educ Health Promot. 2023; 12: 121. doi: 10.4103/jehp.jehp_1008_22. eCollection 2023

Asim, M., Ahmed, ZH., Nichols, AR., Rickman, R., Neiterman, E., Mahmood, A., and Widen. EM. (2021).What stops us from eating: a qualitative investigation of dietary barriers during pregnancy in Punjab, Pakistan. Public Health Nutrition: 25(3), 760–769 doi:10.1017/S1368980021001737

Mohammed, SH., Taye, H., Larijani, B., Esmaillzadeh, A. (2019). Food taboo among pregnant Ethiopian women: magnitude, drivers, and association with anemia. Nutrition Journal 18, Article number: 19 (2019) https://nutritionj.biomedcentral.com/articles/10.1186/s12937-019-0444 doi: 10.1186/s12937-019-0444-4

Laisser, R., Woods, R., Bedwell, C., Kasengele, C., Nsemwa, L., Kimaro, D., Kuzenza, F., Lyangenda, K., Shayo, H., Tuwele, K., Wakasiaka, S., Ringia, P., and Lavender, T., (2021). The tipping point of antenatal engagement. ScienceDirect Sexual & Reproductive Healthcare journal homepage: 10.1016/j.srhc.2021.100673

Angraini, DI., Sulastri, D., Hardisman, H., and Yusrawati, Y., (2023). Angraini Model as Effort to Early Detection of Chronic ergy Deficiency in Pregnancy. KEMAS 19 (1) (2023) 102-112 Jurnal Kesehatan Masyarakat http://journal.unnes.ac.id/nju/index.php/kemas pISSN 1858-1196 eISSN 2355-3596

Workneh, F., Tsegaye, S., Amanuel, H., Eglovitch, M., Shifraw, T., Shiferie, F., Tadesse, AW., Worku, A., Isanaka, S., Lee, ACC., Berhane, Y. 2023. Dietary Perspectives and Practices during Pregnancy in Rural Amhara Region of Ethiopia: An Exploratory Qualitative Study. Current Developments in Nutrition. Volume 7, Issue 6, June 2023, 100079. https://www.sciencedirect.com/science/article/pii/S2475299123212524

Dalaba, MA., Nonterah, EA., Chatio, ST., Adoctor, JK., Watson, D., Barker, M., Ward, KA., Debpuur, C (2021).Culture and community perceptions on diet for maternal and child health: a qualitative study in rural northern Ghana. BMC Nutrition volume 7, Article number: 36 https://bmcnutr.biomedcentral.com/articles/10.1186/s40795-021-00439-x

Alehegn, MA., Fanta, TK., and Ayalew, AF. (2019). Exploring maternal nutrition counseling provided by health professionals during antenatal care follow-up: a qualitative study in Addis Ababa, Ethiopia-2019. BMC Nutrition 7:20 10.1186/s40795-021-00427-1

Darmawati Darmawati., Siregar, TN., Hajjul, K., Tahlil, T.,(2020). Exploring Indonesian mothers’ perspectives on anemia during pregnancy. Enferm Clin (Engl Ed) 2020 Dec 28:S1130-8621(20)30551-9. doi: 10.1016/j.enfcli.2020.11.002

Darmawati Darmawati., Siregar, TN., Hajjul, K., Tahlil, T.,(2020). Acehnese Cultural Leaders’ Perspective on Anemia in Pregnant Women. Hindawi Advances in Public Health., Article ID 8710254, 6 pages 10.1155/2020/8710254

Darmawati Darmawati., Siregar, TN., Hajjul, K., Husna, C.,(2022). Husband’s Perception on Anemia among Pregnant Women based on Cultural Perspective: A Qualitative Study. Macedonia Journal Of Medical Sciences. Vol. 10 No. G (2022):Nursing doi: 10.3889/oamjms.2022.7617

Maykondo, BK., Horwood, C., Haskins, L., Mapumulo, L., MA. Kilola, BM., Mokanisa, MB., Hatloy, A., John, VM. Bitadi, PMB. (2022).A qualitative study to explore dietary knowledge,beliefs, and practices among pregnant women in a rural health zone in the Democratic Republic of Congo. Journal of Health, Population and Nutrition 10.1186/s41043-022-00333-7

Amare, E., Tura, AK., Semahegn, A., and Teji Roba., KT. Food taboos among pregnant women and associated factors in eastern Ethiopia (2022).SAGE Open Medicine Volume 10: 1-11.sagepub.com/journals-permissions DOI: 10.1177/20503121221133935

Ramulondi, M., Wet, H., and Rosemary Ntul, NR. Traditional food taboos and practices during pregnancy, postpartum recovery, and infant care of Zulu women in northern KwaZulu-Nata (2021). Journal of Ethnobiology and Ethnomedicine 17:15 10.1186/s13002-021-00451-2

Rekha V., Manan R., (2019) Coresidence with mother-in-law and maternal anemia in rural India. Social Science & Medicine Volume 226 April 2019 Pages 37-46. https://www.sciencedirect.com/science/article/abs/pii/S0277953619300942

Agbozo. F., Abubakari. A., Der, J., Jahn.(2020).Maternal Dietary Intakes, Red Blood Cell Indices and Risk for Anemia in the First, Second and Third Trimesters of Pregnancy and at Predelivery. Nutrients, 12(3),777; https://www.mdpi.com/2072-6643/12/3/777

Sah, SK., Sunuwar, DR., Baral, JR., Singh, DR., Chaudhary, NK., Gurung, G., (2022). Maternal hemoglobin and risk of low birth weight: A hospital-based cross-sectional study in Nepal.Heliyon. Volume 8, Issue 12, December 2022, e12174. https://www.sciencedirect.com/science/article/pii/S2405844022034624

Citrakesumasari, Dwi, S., Suriah, Bohari, Mesra. R.(2020). Culture-based educate innovation for nutritional status. Enfermería Clínica Volume 30, Supplement 4, June 2020, Pages 9-12. 10.1016/j.enfcli.2019.10.032

Nahrisah, P., Somrongthong, R., Viriyautsahakul, N., Viwattanakulvanid, P., and Plianbangchang, S. (2020). Effect of Integrated Pictorial Handbook Education and Counseling on Improving Anemia Status, Knowledge, Food Intake, and Iron Tablet Compliance Among Anemic Pregnant Women in Indonesia. J Multidiscip Healthc. 2020; 13: 43–52. Published online 2020 Jan 15. doi: 10.2147/JMDH.S213550

Papandreou, P., Vezou, C., Gioxari, A., Kaliora, AC., Skouroliakou. (2023) Improving Adherence to the Mediterranean Dietin Early Pregnancy Using a Clinical Decision Support System (CDSS); A Randomised Controlled Clinical Trial. Nutrients. 2023 Jan; 15(2): 432. Published online 2023 Jan 14. doi: 10.3390/nu15020432. PMCID: PMC9866975 PMID: 3667830. https://www.ncbi.nlm.nih.gov/pmc/articles/PMC9866975/

